## Supplementary material for "Modelling addition and replacement mechanisms of plasmid-based beta-lactam resistant *E. coli* infections": All supporting information

**This PDF file includes:**

Supplementary text 1 to text 4
Figures S1 to S4
Tables S1 to S7
SI References

**Supplementary Information Text**

**Supplementary text 1. Differential equations of the mathematical model.**

The differential equations for the three sub-populations in total are:

$$\frac{dh}{dt} = \alpha * o + v * f - \mu * o$$

[A]

$$\frac{df}{dt} = \mu * o - (\gamma + v) * f$$

[B]

$$\frac{do}{dt} = \gamma * f - \alpha * o$$

[C]

The differential equations for the hospital population are:

$$\frac{dy_{rh}}{dt} = vy_{rf} + \alpha y_{ro} + \lambda_{rr}y_{rrh} - \mu y_{rh} - \beta_{rh} * \left(\frac{y_{rrh}}{sizeh}\right) * y_{rh} - \beta_{rsh} * \left(\frac{\frac{y_{rsh}}{2}}{sizeh}\right) * y_{rh} - \beta_{rrh} * \left(\frac{y_{rrh}}{sizeh}\right) * y_{rh} - \beta_{srh} * \left(\frac{\frac{y_{rsh}}{2}}{sizeh}\right) * y_{rh} - \beta_{sh} * \left(\frac{y_{sh}}{sizeh}\right) * y_{rh} - \beta_{ssh} * \left(\frac{y_{ssh}}{sizeh}\right) * y_{rh} - \omega_r y_{rh} + \frac{1}{2}\lambda_{rs}y_{rsh} + \varphi * (y_{rsh} * \varepsilon_h)$$

[1]

$$\frac{dy_{rrh}}{dt} = \beta_{rh} * (y_{rh}/sizeh) * y_{rh} + \beta_{srh} * \left(\frac{\frac{y_{rsh}}{2}}{sizeh}\right) * y_{rh} + \beta_{rrh} * (y_{rrh}/sizeh) * y_{rh} + \omega_r y_{rh} + vy_{rrf} + \alpha y_{rro} - (\lambda_{rr} + \mu)y_{rrh}$$

[2]

$$\frac{dy_{sh}}{dt} = vy_{sf} + \alpha y_{s0} + \lambda_{ss}y_{ssh} - \mu y_{sh} - \beta_{sh} * \left(\frac{y_{sh}}{sizeh}\right) * y_{sh} - \beta_{ssh} * \left(\frac{y_{ssh}}{sizeh}\right) * y_{sh} - \beta_{rsh} * \left(\frac{\frac{y_{rsh}}{2}}{sizeh}\right) * y_{sh} - \beta_{rh} * \left(\frac{y_{rh}}{sizeh}\right) * y_{sh} - \beta_{rrh} * \left(\frac{y_{rrh}}{sizeh}\right) * y_{sh} - \beta_{srh} * \left(\frac{\frac{y_{rsh}}{2}}{sizeh}\right) * y_{sh} - \omega_s y_{sh} + \frac{1}{2}\lambda_{rs}y_{rsh} + \varphi * (y_{ssh} * \varepsilon_h)$$

[3]

$$\frac{dy_{ssh}}{dt} = \beta_{ssh} * \left(\frac{y_{ssh}}{sizeh}\right) * y_{sh} + \beta_{sh} * \left(\frac{y_{sh}}{sizeh}\right) * y_{sh} + \beta_{rsh} * \left(\frac{\frac{y_{rsh}}{2}}{sizeh}\right) * y_{sh} + \omega_s y_{sh} + vy_{ssf} + \alpha y_{sso} - (\lambda_{ss} + \mu)y_{ssh} - \varphi * (y_{ssh} * \varepsilon_h)$$

[4]

$$\frac{dy_{rsh}}{dt} = \beta_{rsh}\left(\frac{\frac{y_{rsh}}{2}}{sizeh}\right) * y_{rh} + \beta_{sh} * \left(\frac{y_{sh}}{sizeh}\right) * y_{rh} + \beta_{ssh} * \left(\frac{y_{ssh}}{sizeh}\right) * y_{rh} + \beta_{rh} * \left(\frac{y_{rh}}{sizeh}\right) * y_{sh} + \beta_{rrh} * \left(\frac{y_{rrh}}{sizeh}\right) * y_{sh} + \beta_{srh} * \left(\frac{\frac{y_{rsh}}{2}}{sizeh}\right) * y_{sh} + v y_{rsf} + \alpha y_{rso} - (\lambda_{rs} + \mu) y_{rsh} - \varphi * (y_{rsh} * \varepsilon_h)$$

[5]

The differential equations for the former patients in the community are:

$$\frac{dy_{rf}}{dt} = \mu y_{rh} + \lambda_{rr} y_{rrf} - (v + \gamma) y_{rf} - \beta_{rf} * \left(\frac{y_{rf}}{sizef + sizeo}\right) * y_{rf} - \beta_{rrf} * \left(\frac{y_{rrf}}{sizef + sizeo}\right) * y_{rf} - \beta_{rsf} * \left(\frac{\frac{y_{rsh}}{2}}{sizef + sizeo}\right) * y_{rf} - \beta_{sf} * \left(\frac{y_{sf}}{sizef + sizeo}\right) * y_{rf} - \beta_{ssf} * \left(\frac{y_{ssf}}{sizef + sizeo}\right) * y_{rf} - \beta_{srf} * \left(\frac{\frac{y_{rsh}}{2}}{sizef + sizeo}\right) * y_{rf} - \beta_{ro} * \left(\frac{y_{ro}}{sizef + sizeo}\right) * y_{rf} - \beta_{rro} * \left(\frac{y_{rro}}{sizef + sizeo}\right) * y_{rf} - \beta_{rso} * \left(\frac{\frac{y_{rso}}{2}}{sizef + sizeo}\right) * y_{rf} - \beta_{so} * \left(\frac{y_{so}}{sizef + sizeo}\right) * y_{rf} - \beta_{sso} * \left(\frac{y_{sso}}{sizef + sizeo}\right) * y_{rf} - \beta_{sro} * \left(\frac{\frac{y_{rso}}{2}}{sizef + sizeo}\right) * y_{rf} - \omega_r y_{rf} + \frac{1}{2}\lambda_{rs} y_{rsf} + \varphi * (y_{rsf} * \varepsilon_f)$$

[6]

$$\frac{dy_{rrf}}{dt} = \beta_{rf} * \left(\frac{y_{rf}}{sizef + sizeo}\right) * y_{rf} + \beta_{rrf} * \left(\frac{y_{rrf}}{sizef + sizeo}\right) * y_{rf} + \beta_{srf} * \left(\frac{\frac{y_{rsh}}{2}}{sizef + sizeo}\right) * y_{rf} + \omega_r y_{rf} + \beta_{ro} * \left(\frac{y_{ro}}{sizef + sizeo}\right) * y_{rf} + \beta_{rro} * \left(\frac{y_{rro}}{sizef + sizeo}\right) * y_{rf} + \beta_{sro} * \left(\frac{\frac{y_{rso}}{2}}{sizef + sizeo}\right) * y_{rf} + \omega_r y_{rf} + \mu y_{rrh} - (v + \lambda_{rr} + \gamma) y_{rrf}$$

[7]

$$\frac{dy_{sf}}{dt} = \mu y_{sh} + \lambda_{ss} y_{ssf} - (v + \gamma) y_{sf} - \beta_{rf} * \left(\frac{y_{rf}}{sizef + sizeo}\right) * y_{sf} - \beta_{rrf} * \left(\frac{y_{rrf}}{sizef + sizeo}\right) * y_{sf} - \beta_{srf} * \left(\frac{\frac{y_{rsh}}{2}}{sizef + sizeo}\right) * y_{sf} - \beta_{ro} * \left(\frac{y_{ro}}{sizef + sizeo}\right) * y_{sf} - \beta_{rro} * \left(\frac{y_{rro}}{sizef + sizeo}\right) * y_{sf} - \beta_{sro} * \left(\frac{\frac{y_{rso}}{2}}{sizef + sizeo}\right) * y_{sf} - \beta_{sf} * \left(\frac{y_{sf}}{sizef + sizeo}\right) * y_{sf} - \beta_{ssf} * \left(\frac{y_{ssf}}{sizef + sizeo}\right) * y_{sf} - \beta_{rsf} * \left(\frac{\frac{y_{rsh}}{2}}{sizef + sizeo}\right) * y_{sf} - \beta_{so} * \left(\frac{y_{so}}{sizef + sizeo}\right) * y_{sf} - \beta_{sso} * \left(\frac{y_{sso}}{sizef + sizeo}\right) * y_{sf} - \beta_{rso} * \left(\frac{\frac{y_{rso}}{2}}{sizef + sizeo}\right) * y_{sf} - \omega_s y_{sf} + \frac{1}{2} \lambda_{rs} y_{rsf} + \varphi * (y_{ssf} * \varepsilon_f)$$

[8]

$$\frac{dy_{ssf}}{dt} = \beta_{sf} * \left(\frac{y_{sf}}{sizef + sizeo}\right) * y_{sf} + \beta_{ssf} * \left(\frac{y_{ssf}}{sizef + sizeo}\right) * y_{sf} + \beta_{rsf} * \left(\frac{\frac{y_{rsh}}{2}}{sizef + sizeo}\right) * y_{sf} + \beta_{so} * \left(\frac{y_{so}}{sizef + sizeo}\right) * y_{sf} + \beta_{sso} * \left(\frac{y_{sso}}{sizef + sizeo}\right) * y_{sf} + \beta_{rso} * \left(\frac{\frac{y_{rso}}{2}}{sizef + sizeo}\right) * y_{sf} + \omega_s y_{sf} + \mu y_{ssh} - (v + \lambda_{ss} + \gamma) y_{ssf} - \varphi * (y_{ssf} * \varepsilon_f)$$

[9]

$$\frac{dy_{rsf}}{dt} = \beta_{rf} * \left(\frac{y_{rf}}{sizef + sizeo}\right) * y_{sf} + \beta_{rrf} * \left(\frac{y_{rrf}}{sizef + sizeo}\right) * y_{sf} + \beta_{srf} * \left(\frac{\frac{y_{rsh}}{2}}{sizef + sizeo}\right) * y_{sf} + \beta_{ro} * \left(\frac{y_{ro}}{sizef + sizeo}\right) * y_{sf} + \beta_{rro} * \left(\frac{y_{rro}}{sizef + sizeo}\right) * y_{sf} + \beta_{sro} * \left(\frac{\frac{y_{rso}}{2}}{sizef + sizeo}\right) * y_{sf} + \beta_{sf} * \left(\frac{y_{sf}}{sizef + sizeo}\right) * y_{rf} + \beta_{ssf} * \left(\frac{y_{ssf}}{sizef + sizeo}\right) * y_{rf} - \beta_{rsf} * \left(\frac{\frac{y_{rsh}}{2}}{sizef + sizeo}\right) * y_{rf} + \beta_{rso} * \left(\frac{\frac{y_{rso}}{2}}{sizef + sizeo}\right) * y_{rf} + \beta_{so} * \left(\frac{y_{so}}{sizef + sizeo}\right) * y_{rf} + \beta_{sso} * \left(\frac{y_{sso}}{sizef + sizeo}\right) * y_{rf} + \mu y_{rsh} - (v + \lambda_{rs} + \gamma) y_{rsf} - \varphi * (y_{rsf} * \varepsilon_f)$$

[10]

The differential equations for the open population in the community are:

$$\frac{dy_{ro}}{dt} = \gamma y_{rf} + \lambda_{rr} y_{rro} - \alpha y_{ro} - \beta_{rf} * \left(\frac{y_{rf}}{sizef + sizeo}\right) * y_{ro} - \beta_{rrf} * \left(\frac{y_{rrf}}{sizef + sizeo}\right) * y_{ro} - \beta_{rsf} * \left(\frac{\frac{y_{rsh}}{2}}{sizef + sizeo}\right) * y_{ro} - \beta_{sf} * \left(\frac{y_{sf}}{sizef + sizeo}\right) * y_{ro} - \beta_{ssf} * \left(\frac{y_{ssf}}{sizef + sizeo}\right) * y_{ro} - \beta_{srf} * \left(\frac{\frac{y_{rsh}}{2}}{sizef + sizeo}\right) * y_{ro} - \beta_{ro} * \left(\frac{y_{ro}}{sizef + sizeo}\right) * y_{ro} - \beta_{rro} * \left(\frac{y_{rro}}{sizef + sizeo}\right) * y_{ro} - \beta_{rso} * \left(\frac{\frac{y_{rso}}{2}}{sizef + sizeo}\right) * y_{ro} - \beta_{so} * \left(\frac{y_{so}}{sizef + sizeo}\right) * y_{ro} - \beta_{sso} * \left(\frac{y_{sso}}{sizef + sizeo}\right) * y_{ro} - \beta_{sro} * \left(\frac{\frac{y_{rso}}{2}}{sizef + sizeo}\right) * y_{ro} - \omega_r y_{ro} + \frac{1}{2}\lambda_{rs} y_{rso} + \varphi * (y_{rso} * \varepsilon_o)$$

[11]

$$\frac{dy_{rro}}{dt} = \beta_{rf} * \left(\frac{y_{rf}}{sizef + sizeo}\right) * y_{ro} + \beta_{rrf} * \left(\frac{y_{rrf}}{sizef + sizeo}\right) * y_{ro} + \beta_{srf} * \left(\frac{\frac{y_{rsh}}{2}}{sizef + sizeo}\right) * y_{ro} + \beta_{ro} * \left(\frac{y_{ro}}{sizef + sizeo}\right) * y_{ro} + \beta_{rro} * \left(\frac{y_{rro}}{sizef + sizeo}\right) * y_{ro} + \beta_{sro} * \left(\frac{\frac{y_{rso}}{2}}{sizef + sizeo}\right) * y_{ro} + \omega_r y_{ro} + \gamma y_{rrf} - (\alpha + \lambda_{rr}) yr_{rro}$$

[12]

$$\frac{dy_{so}}{dt} = \gamma y_{sf} + \lambda_{ss} y_{sso} - \alpha y_{so} - \beta_{rf} * \left(\frac{y_{rf}}{sizef + sizeo}\right) * y_{so} - \beta_{rrf} * \left(\frac{y_{rrf}}{sizef + sizeo}\right) * y_{so} - \beta_{srf} * \left(\frac{\frac{y_{rsh}}{2}}{sizef + sizeo}\right) * y_{so} - \beta_{ro} * \left(\frac{y_{ro}}{sizef + sizeo}\right) * y_{so} - \beta_{rro} * \left(\frac{y_{rro}}{sizef + sizeo}\right) * y_{so} - \beta_{sro} * \left(\frac{\frac{y_{rso}}{2}}{sizef + sizeo}\right) * y_{so} - \beta_{sf} * \left(\frac{y_{sf}}{sizef + sizeo}\right) * y_{so} - \beta_{ssf} * \left(\frac{y_{ssf}}{sizef + sizeo}\right) * y_{so} - \beta_{rsf} * \left(\frac{\frac{y_{rsh}}{2}}{sizef + sizeo}\right) * y_{so} - \beta_{so} * \left(\frac{y_{so}}{sizef + sizeo}\right) * y_{so} - \beta_{sso} * \left(\frac{y_{sso}}{sizef + sizeo}\right) * y_{so} - \beta_{rso} * \left(\frac{\frac{y_{rso}}{2}}{sizef + sizeo}\right) * y_{so} - \omega_s y_{so} + \frac{1}{2}\lambda_{rs} y_{rso} + \varphi * (y_{sso} * \varepsilon_o)$$

[13]

$$\frac{dy_{sso}}{dt} = \beta_{sf} * \left(\frac{y_{sf}}{sizef + sizeo}\right) * y_{so} + \beta_{ssf} * \left(\frac{y_{ssf}}{sizef + sizeo}\right) * y_{so} + \beta_{rsf} * \left(\frac{\frac{y_{rsh}}{2}}{sizef + sizeo}\right) * y_{so} + \beta_{so} * \left(\frac{y_{so}}{sizef + sizeo}\right) * y_{so} + \beta_{sso} * \left(\frac{y_{sso}}{sizef + sizeo}\right) * y_{so} + \beta_{rso} * \left(\frac{\frac{y_{rso}}{2}}{sizef + sizeo}\right) * y_{so} + \omega_s y_{so} + \gamma y_{ssf} - (\alpha + \lambda_{ss}) y_{sso} - \varphi * (y_{sso} * \varepsilon_o)$$

[14]

$$\frac{dy_{rso}}{dt} = \beta_{rf} * \left(\frac{y_{rf}}{sizef + sizeo}\right) * y_{so} + \beta_{rrf} * \left(\frac{y_{rrf}}{sizef + sizeo}\right) * y_{so} + \beta_{srf} * \left(\frac{\frac{y_{rsh}}{2}}{sizef + sizeo}\right) * y_{so} + \beta_{ro} * \left(\frac{y_{ro}}{sizef + sizeo}\right) * y_{so} + \beta_{rro} * \left(\frac{y_{rro}}{sizef + sizeo}\right) * y_{so} + \beta_{sro} * \left(\frac{\frac{y_{rso}}{2}}{sizef + sizeo}\right) * y_{so} + \beta_{sf} * \left(\frac{y_{sf}}{sizef + sizeo}\right) * y_{ro} + \beta_{ssf} * \left(\frac{y_{ssf}}{sizef + sizeo}\right) * y_{ro} + \beta_{rsf} * \left(\frac{\frac{y_{rsh}}{2}}{sizef + sizeo}\right) * y_{ro} + \beta_{rso} * \left(\frac{\frac{y_{rso}}{2}}{sizef + sizeo}\right) * y_{ro} + \beta_{so} * \left(\frac{y_{so}}{sizef + sizeo}\right) * y_{ro} + \beta_{sso} * \left(\frac{y_{sso}}{sizef + sizeo}\right) * y_{ro} + \gamma y_{rsf} - (\alpha + \lambda_{rs}) y_{rso} - \varphi * (y_{rso} * \varepsilon_o)$$

[15]

The code in Wolfram Mathematica 12.1 is available on https://github.com/NoorGo/AdditionandReplacement

**Supplementary text 2. Infection rate calculations**

In 2017, 159,619 infections with *E. Coli* were cultured in 34 laboratories the Netherlands (1). Of these, 57% occurred at the general practitioner and 15% in outpatients. Thus, in total roughly 72% of the *E. coli* infections that were severe enough to pose the need to be cultured in 2017 occurred in the community, which are 114,925 infections.

The Netherlands had 17,08 million inhabitants in 2017. In our model, 0.177% of the people is in the hospital, Thus, this would mean that in 2017 (1-0.00177)*17.08= 17.05 people were in the open population. Based on the number of infections in de Greeff & Mouton, 2018, the infection rate per year per person is 114,925/170,497684 =$6.7406x10^{-4}$ and the infection rate per day per person is $6.7406x10^{-4}$/365=$1.846745046839801x10^{-6}$. However, the number of infections reported in de Greeff & Mouton, 2018 is an underrepresentation of all infections in the Netherlands, as not all Dutch medical microbiological laboratories were included in the analyses of this report.

To arrive at a total of 72% of the infections occurring in the open population (as observed in the Netherlands), we have to adjust this rate. Based on the previously research transmission rates we reported in Table S2, 684 infections would occur in the hospital. If this is 28% of the total number of infections, then 1,758 infections should occur in the open population. As 1,342 infections occur in the former patient population based on the estimates in Table S2, 416 should occur in the open population. With a rate of $1.846745046839801x10^{-6,}$ only 66 infections would occur in the open population which is as expected an underestimate. Dividing 416 by 66 gives 6.22589616612008, and multiplying the infection rate of the open population with this number gives a total number of 416 infections in the open population. To arrive at this number, we have to multiply the infection rate of $1.846745046839801x10^{-6}$ with 6.22589616612008.

**Supplementary text 3. Calculations admission and readmission rates**

In 2017, the Netherlands had a total number of 37,753 hospital beds (2). Assuming an occupancy of 80%, this would mean that per day there were 0.8*37,753= 30,202 people in the hospital. Since there were 17,08 million inhabitants in 2017, a proportion of 30,202/17,0800000 = 0.00177 of the Dutch population is in the hospital. Per 100,000 inhabitants this is 177 people per day that stay in the hospital.

To arrive at this number, the estimates of Cooper et al. (3) have to be multiplied with a fraction of 0.333183. We arrived at this number by solving equation A-C, given that the hospital population is 177. The admission and readmission rate will stay proportional to the estimates in Cooper et al. (3) and now become 0.00063*0.333183=0.00020990529 and 0.0057*0.333183 =0.0018991431, which indeed results in 177 people in the hospital per day per 100,000.

**Supplementary text 4. External plasmid transfer**

Plasmid transfer may occur from other resistant pathogens to *E. coli.* In this mechanism, we model plasmid transfer from other sources. We add a route from SS to SR and from SS to RR. In addition, we assume that when an individual colonized with just an S strain acquires a resistant plasmid, the individual moves to SR rather than R because not all susceptible *E. coli* the person is colonized with will become resistant immediately. This results in the compartmental model in Figure S1.

We started with a rate of 1% and increased this rate with 10% each time, thus ending with a rate of 2% (a 100% increase of 1%). We also modelled a rate 0·1% and increased it with 10%, ending with a rate of 0.2%. Both scenarios caused for total replacement of the susceptible strain after 50 years.

**Supplementary text 5. Calculations of increased virulence**
Table S8 shows the observed incidence density of Enterobacteriaceae in the paper of Ammerlaan et al. (4) and the expected incidence density of Enterobacteriaceae if the ARB Enterobacteriaceae would have grown equally.

An increase of virulence of 10.5 times is needed to arrive at 10·12 instead of 0.97. This is estimated as 10.2 / 0.97.

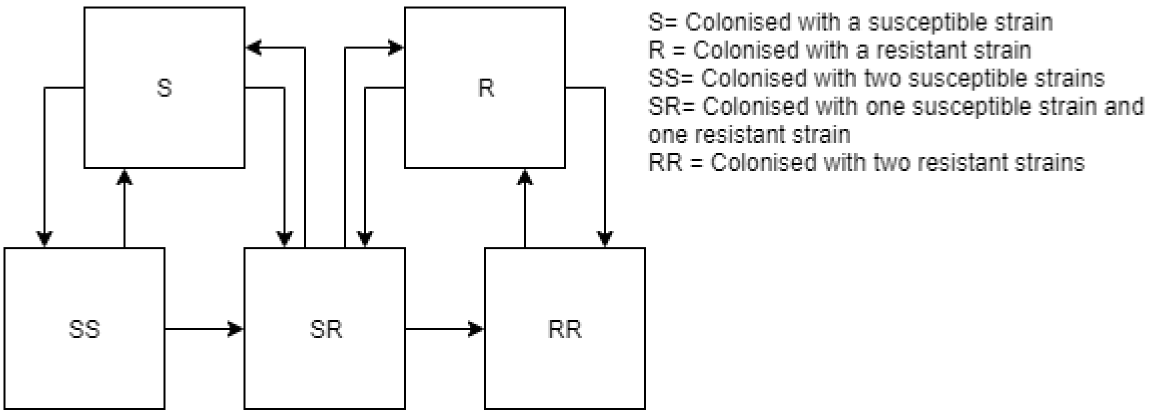


**Fig. S1.** Compartmental model of E. coli with plasmid transfer from other sources

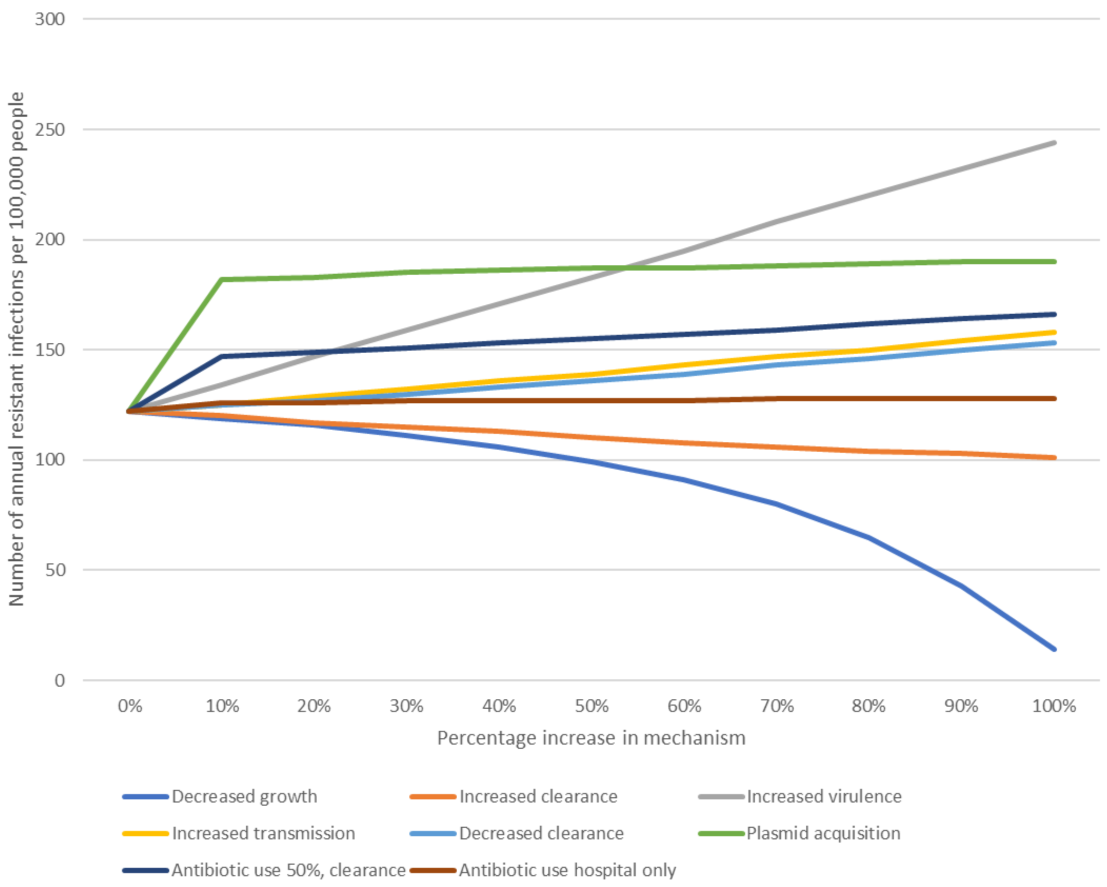


**Fig. S2.** Annual ESBL *E. coli* infections per 100,000 people per mechanism in 10 years of time

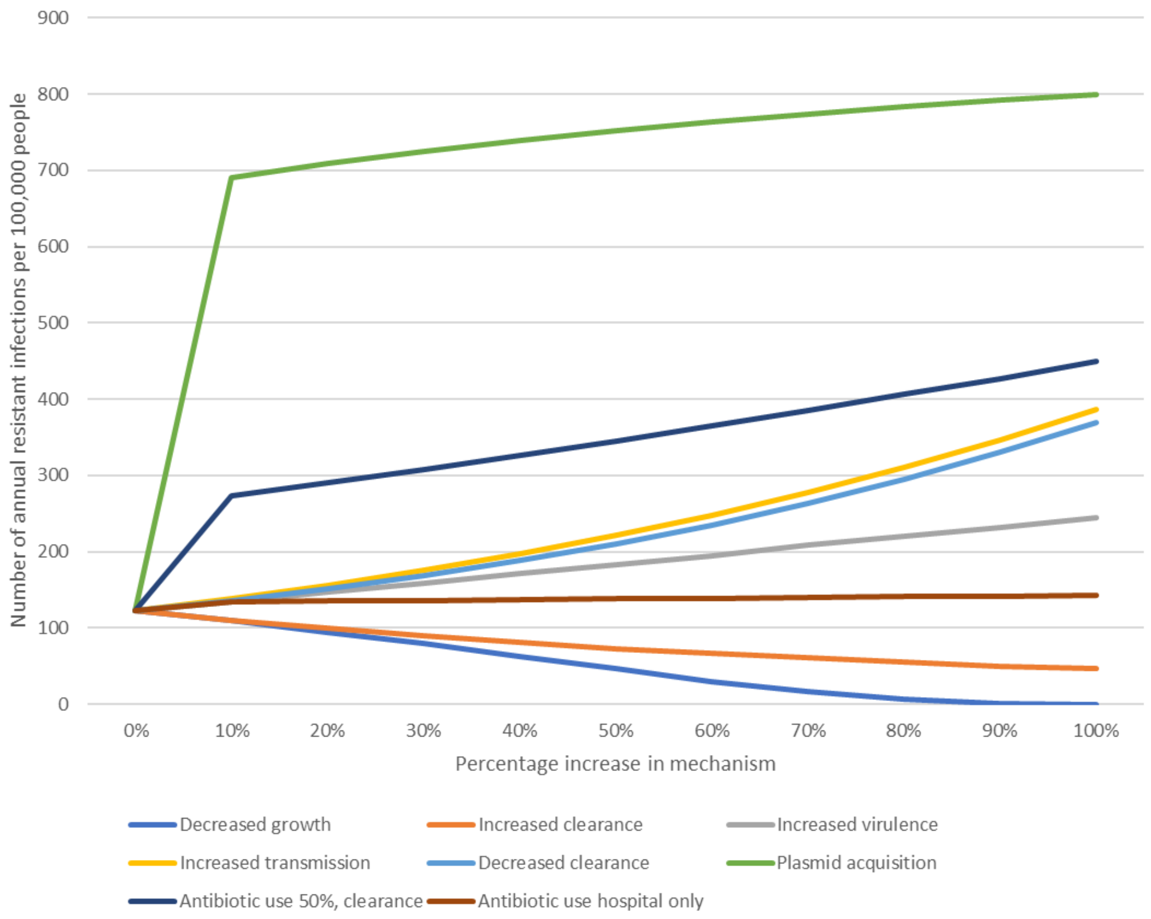


**Fig. S3.** Annual ESBL *E. coli* infections per 100,000 people per mechanism in in 50 years of time

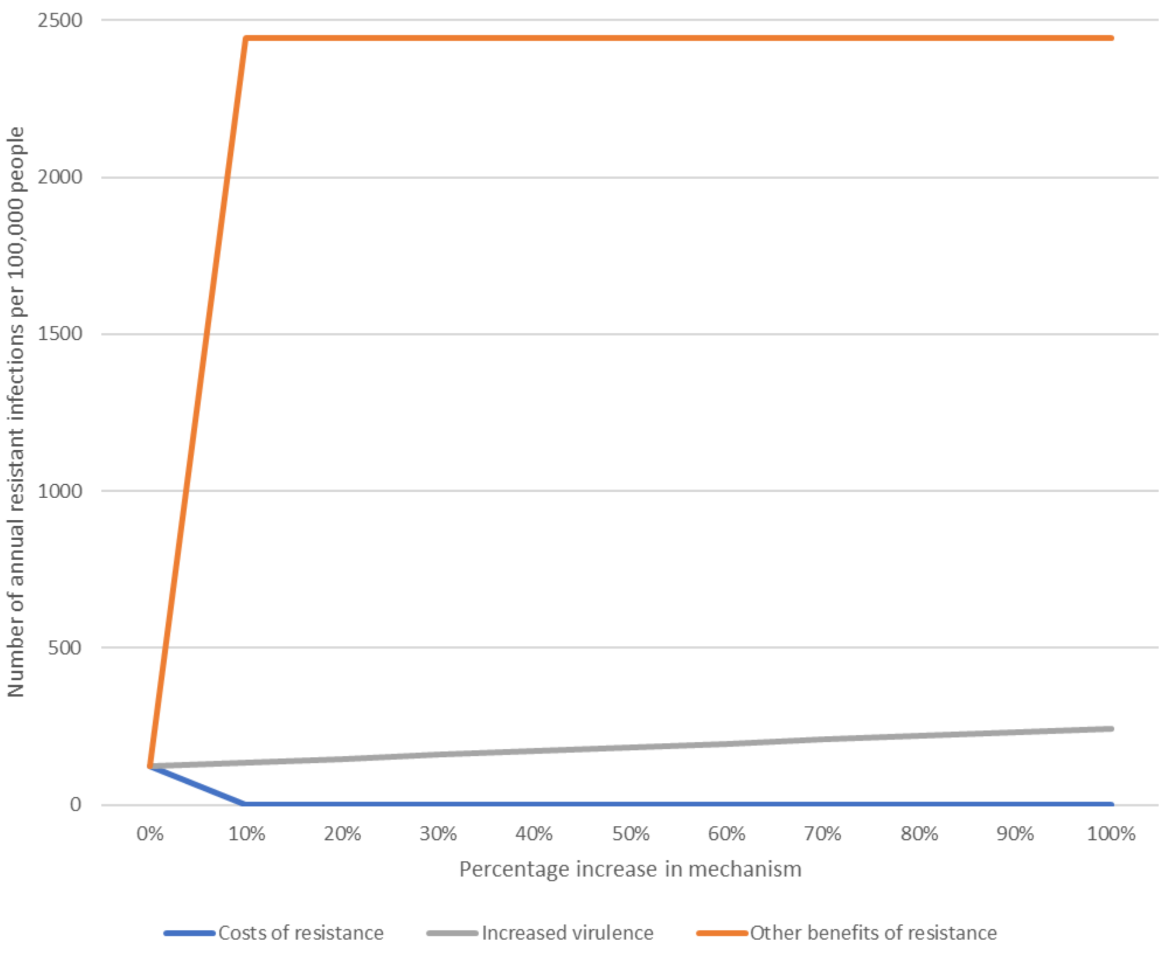


**Fig. S4.** Annual ESBL E. coli infections per 100,000 people per mechanism when letting time run to infinity

**Table S1.** Annual prevalence of ESBL in E. coli bacteraemia in 24 Dutch hospitals

| Year | Tested samples | ESBL | Prevalence of ESBL (%) | 95% CI |
|---|---|---|---|---|
| 2014 | 2675 | 144 | 5.38 | 4·59 – 6·31 |
| 2015 | 2730 | 147 | 5.38 | 4·60 – 6·30 |
| 2016 | 2800 | 166 | 5.93 | 5·11 – 6·87 |
| 2017 | 2842 | 153 | 5.38 | 4·61 – 6·28 |
| 2018 | 3037 | 168 | 5.53 | 4·77 – 6·40 |

*ESBL* = extended spectrum beta-lactamases producing, *CI = confidence interval*

**Table S2.** Scenarios and mechanisms studied in this paper

| | | | Rate per day | | |
|---|---|---|---|---|---|
| Scenario | Mechanism | Adjusted path | Hospitalized patients | Former patients | Open population |
| Fitness cost | Increased clearance | Increases RR to R & SR to S | 0.0028[a]-0.0056 | 0.0028[a]-0.0056 | 0.0028[a]-0.0056 |
| | Decreased growth | Decreases R to RR | 0.0357[a]-0.00357 | 0.0357[a]-0.00357 | 0.0357[a]-0.00357 |
| Benefit | Increased virulence | Increases probability resistant infections | 0.106[a]-0.212 | 0.053[a]-0.106 | 1.15^10$^{-5}$[a]-2.30^10$^{-5}$ |
| | Increased transmission | Increases S to SR & R to RR | 0.0078[a]-0.0156 | 0.0053[a]-0.0106 | 0.00195[a]-0.0039 |
| | Decreased clearance | Decreases RR to R & SR to S | 0.0028[a]-0.00028 | 0.0028[a]-0.00028 | 0.0028[a]-0.00028 |
| | Plasmid transfer | Creates path SR to RR | 0·010[a]-0·02 | 0·010[a]-0·02 | 0·010[a]-0·02 |
| | Antibiotic use | Increases SS to S & SR to R | 0.064[a]-0.128 | 0.032[a]– 0.065 | 0.001-0.002 |
| | Hospital antibiotic use | Increases SS to S & SR to R | 0.34[a]-0.68 | 0[a] | 0[a] |
| Mixed scenario | 20% increased hospital transmission | Increases S to SR & R to RR | 0.0094 | | |
| | 20% increased clearance in open population | Increases RR to R & SR to S | 0.0034 | | |
| Double benefit scenario | 20% increased virulence | Inc. probability resistant infections | 0.127 | 0.064 | 1.38^10$^{-5}$ |
| | 20% increased transmission | Increases S to SR & R to RR | 0.0094 | 0.0064 | 0.00234 |

[a] Starting value, the same value as for the susceptible variant

**Table S3.** In 50 years, the number of resistant and susceptible infections per 100,000 inhabitants under different scenarios

| Infections | Neutral | Mixed | Double benefit |
|---|---|---|---|
| Resistant (ARB) | 122 | 100 | 702 |
| Susceptible (non-ARB) | 2320 | 2343 | 1857 |
| Total | 2443 | 2443 | 2560 |

**Table S4.** Parameters used in the model

| Parameter[1] | Symbol | Hospital $_h$[2] | Former patients $_f$[2] | Open population $_o$[2] |
|---|---|---|---|---|
| Transmission | $\beta$ | 0·0078(5) | 0·0053(6) | 0·00195 ($\beta_h$ / 4) |
| Clearance rate | $\lambda$ | 0·0028(7) | 0·0028(7) | 0·0028(7) |
| Infection rate hospital | $\pi$ | 0·106(8) | 0·053 ($\pi_h$ /2) | $1{\cdot}14874{*}10^{-5}$ (Supplementary text 2, own calculations on data of de Greeff & Mouton (1) |
| Admission rate | $\alpha$ | | | 0·00020991 (Adjusted from Cooper et al. (3) based on Dutch data. Supplementary text 3) |
| Readmission rate | $v$ | | 0·0018991 (Adjusted from Cooper et al. (3) based on Dutch data. Supplementary text 3) | |
| Discharge rate | $\mu$ | 0·125(3) | | |
| Rate to open population | $\gamma$ | | 0·03(3) | |
| Rate of growing to high density | $\Omega$ | 0·0357(9) | 0·0357(9) | 0·0357(9) |
| Proportion receiving antibiotics* | $\varepsilon$ | | 0 | |
| Proportion cleared of susceptible strain due to antibiotics* | $\varphi$ | 0.5 | 0.5 | 0.5 |
| Plasmid transfer* | $\vartheta$ | 0·01 | 0·01 | 0·01 |
| External plasmid transfer* | $\vartheta_2$ | 0·001 | 0·001 | 0·001 |

* Value is 0 in the neutral model

1 For each rate, subscripts $_s$ (susceptible), $_{ss}$ (high density susceptible), $_{sr}$ (resistant and susceptible), $_r$ (resistant) and $_{rr}$ (high density resistant) indicated the rate attributable to those with that specific colonisation state.

2 Subscripts $_h$ (hospital), $_f$ (former patient) and $_o$ (open population) are used to indicate each population.

**Table S5.** In 10 years, the number of infections per 100,000 people per year for each mechanism

| | | Number of infections per 100,000 per year | | | | | | | | | | |
|---|---|---|---|---|---|---|---|---|---|---|---|---|
| | | Percentage of change in characteristic | | | | | | | | | | |
| Altered characteristic | | 0 | 10 | 20 | 30 | 40 | 50 | 60 | 70 | 80 | 90 | 100 |
| Increased clearance | R | 122 | 120 | 117 | 115 | 113 | 110 | 108 | 106 | 104 | 103 | 101 |
| | S | 2320 | 2323 | 2325 | 2328 | 2330 | 2332 | 2334 | 2336 | 2338 | 2340 | 2342 |
| | T | 2443 | 2443 | 2443 | 2443 | 2443 | 2443 | 2443 | 2443 | 2443 | 2443 | 2443 |
| Decreased growth | R | 122 | 119 | 116 | 111 | 106 | 99 | 91 | 80 | 65 | 43 | 14 |
| | S | 2320 | 2320 | 2323 | 2327 | 2331 | 2337 | 2343 | 2352 | 2378 | 2399 | 2429 |
| | T | 2443 | 2443 | 2443 | 2443 | 2443 | 2443 | 2443 | 2443 | 2443 | 2443 | 2443 |
| Increased virulence | R | 122 | 134 | 147 | 159 | 171 | 183 | 195 | 208 | 220 | 232 | 244 |
| | S | 2320 | 2320 | 2320 | 2320 | 2320 | 2320 | 2320 | 2320 | 2320 | 2320 | 2320 |
| | T | 2443 | 2455 | 2467 | 2479 | 2491 | 2504 | 2516 | 2528 | 2540 | 2552 | 2565 |
| Increased transmission | R | 122 | 125 | 129 | 132 | 136 | 139 | 143 | 147 | 150 | 154 | 158 |
| | S | 2320 | 2317 | 2314 | 2311 | 2307 | 2303 | 2300 | 2296 | 2292 | 2288 | 2284 |
| | T | 2443 | 2443 | 2443 | 2443 | 2443 | 2443 | 2443 | 2443 | 2443 | 2443 | 2443 |
| Decreased clearance | R | 122 | 125 | 127 | 130 | 133 | 136 | 139 | 143 | 146 | 150 | 153 |
| | S | 2320 | 2318 | 2315 | 2312 | 2309 | 2306 | 2303 | 2300 | 2296 | 2293 | 2289 |
| | T | 2443 | 2443 | 2443 | 2443 | 2443 | 2443 | 2443 | 2443 | 2443 | 2443 | 2443 |
| Plasmid acquisition | R | 122 | 182 | 183 | 185 | 186 | 187 | 187 | 188 | 189 | 190 | 190 |
| | S | 2320 | 2261 | 2259 | 2258 | 2257 | 2256 | 2255 | 2254 | 2254 | 2253 | 2252 |
| | T | 2443 | 2443 | 2443 | 2443 | 2443 | 2443 | 2443 | 2443 | 2443 | 2443 | 2443 |
| Antibiotic use, 50% clearance | R | 122 | 147 | 149 | 151 | 153 | 155 | 157 | 159 | 162 | 164 | 166 |
| | S | 2320 | 2296 | 2292 | 2291 | 2289 | 2287 | 2285 | 2283 | 2281 | 2279 | 2277 |
| | T | 2443 | 2443 | 2443 | 2443 | 2443 | 2443 | 2443 | 2443 | 2443 | 2443 | 2443 |
| Antibiotic use in hospital only | R | 122 | 126 | 126 | 127 | 127 | 127 | 127 | 128 | 128 | 128 | 128 |
| | S | 2320 | 2317 | 2316 | 2316 | 2316 | 2315 | 2315 | 2315 | 2315 | 2314 | 2314 |
| | T | 2443 | 2443 | 2443 | 2443 | 2443 | 2443 | 2443 | 2443 | 2443 | 2443 | 2443 |

*R = resistant, S = susceptible, T= total*

**Table S6.** In 50 years, the number of infections per 100,000 people per year for each mechanism

| | | Number of infections per 100,000 per year | | | | | | | | | | |
|---|---|---|---|---|---|---|---|---|---|---|---|---|
| | | Percentage of change in characteristic | | | | | | | | | | |
| Altered characteristic | | 0 | 10 | 20 | 30 | 40 | 50 | 60 | 70 | 80 | 90 | 100 |
| Increased clearance | R | 122 | 110 | 99 | 90 | 81 | 73 | 67 | 61 | 55 | 50 | 46 |
| | S | 2320 | 2333 | 2343 | 2353 | 2361 | 2369 | 2376 | 2382 | 2387 | 2392 | 2397 |
| | T | 2443 | 2443 | 2443 | 2443 | 2443 | 2443 | 2443 | 2443 | 2443 | 2443 | 2443 |
| Decreased growth | R | 122 | 109 | 94 | 79 | 62 | 46 | 30 | 16 | 6 | 1 | 0 |
| | S | 2320 | 2334 | 2349 | 2364 | 2380 | 2397 | 2413 | 2427 | 2437 | 2442 | 2443 |
| | T | 2443 | 2443 | 2443 | 2443 | 2443 | 2443 | 2443 | 2443 | 2443 | 2443 | 2443 |
| Increased virulence | R | 122 | 134 | 147 | 159 | 171 | 183 | 195 | 208 | 220 | 232 | 244 |
| | S | 2320 | 2320 | 2320 | 2320 | 2320 | 2320 | 2320 | 2320 | 2320 | 2320 | 2320 |
| | T | 2443 | 2455 | 2467 | 2479 | 2491 | 2504 | 2516 | 2528 | 2540 | 2553 | 2565 |
| Increased transmission | R | 122 | 138 | 155 | 175 | 197 | 221 | 248 | 278 | 311 | 347 | 386 |
| | S | 2320 | 2305 | 2287 | 2268 | 2246 | 2221 | 2194 | 2164 | 2132 | 2096 | 2056 |
| | T | 2443 | 2443 | 2443 | 2443 | 2443 | 2443 | 2443 | 2443 | 2443 | 2443 | 2443 |
| Decreased clearance | R | 122 | 136 | 151 | 168 | 188 | 210 | 235 | 263 | 294 | 330 | 369 |
| | S | 2320 | 2307 | 2291 | 2274 | 2255 | 2233 | 2208 | 2180 | 2148 | 2113 | 2073 |
| | T | 2443 | 2443 | 2443 | 2443 | 2443 | 2443 | 2443 | 2443 | 2443 | 2443 | 2443 |
| Plasmid acquisition | R | 122 | 691 | 709 | 725 | 739 | 752 | 763 | 774 | 783 | 792 | 800 |
| | S | 2320 | 1752 | 1734 | 1718 | 1704 | 1691 | 1679 | 1669 | 1659 | 1651 | 1643 |
| | T | 2443 | 2443 | 2443 | 2443 | 2443 | 2443 | 2443 | 2443 | 2443 | 2443 | 2443 |
| Antibiotic use, 50% clearance | R | 122 | 273 | 290 | 308 | 326 | 345 | 365 | 385 | 406 | 427 | 450 |
| | S | 2320 | 2170 | 2153 | 3135 | 2116 | 2097 | 2078 | 2057 | 2036 | 2015 | 1993 |
| | T | 2443 | 2443 | 2443 | 2443 | 2443 | 2443 | 2443 | 2443 | 2443 | 2443 | 2443 |
| Antibiotic use in hospital only | R | 122 | 134 | 135 | 136 | 137 | 138 | 139 | 140 | 141 | 141 | 142 |
| | S | 2320 | 2309 | 2308 | 2307 | 2306 | 2305 | 2304 | 2303 | 2302 | 2301 | 2300 |
| | T | 2443 | 2443 | 2443 | 2443 | 2443 | 2443 | 2443 | 2443 | 2443 | 2443 | 2443 |

*R = resistant, S = susceptible, T= total*

**Table S7.** Letting time run to infinity, the number of infections per 100,000 people per year for each mechanism

| | | Number of infections per 100,000 per year | | | | | | | | | | |
|---|---|---|---|---|---|---|---|---|---|---|---|---|
| | | Percentage of change in characteristic | | | | | | | | | | |
| Altered characteristic | | 0 | 10 | 20 | 30 | 40 | 50 | 60 | 70 | 80 | 90 | 100 |
| Increased clearance | R | 122 | 0 | 0 | 0 | 0 | 0 | 0 | 0 | 0 | 0 | 0 |
| | S | 2320 | 2443 | 2443 | 2443 | 2443 | 2443 | 2443 | 2443 | 2443 | 2443 | 2443 |
| | T | 2443 | 2443 | 2443 | 2443 | 2443 | 2443 | 2443 | 2443 | 2443 | 2443 | 2443 |
| Decreased growth | R | 122 | 0 | 0 | 0 | 0 | 0 | 0 | 0 | 0 | 0 | 0 |
| | S | 2320 | 2443 | 2443 | 2443 | 2443 | 2443 | 2443 | 2443 | 2443 | 2443 | 2443 |
| | T | 2443 | 2443 | 2443 | 2443 | 2443 | 2443 | 2443 | 2443 | 2443 | 2443 | 2443 |
| Increased virulence | R | 122 | 134 | 147 | 159 | 171 | 183 | 195 | 208 | 220 | 232 | 244 |
| | S | 2320 | 2320 | 2320 | 2320 | 2320 | 2320 | 2320 | 2320 | 2320 | 2320 | 2320 |
| | T | 2443 | 2455 | 2467 | 2479 | 2491 | 2504 | 2516 | 2528 | 2540 | 2552 | 2565 |
| Increased transmission | R | 122 | 2443 | 2443 | 2443 | 2443 | 2443 | 2443 | 2443 | 2443 | 2443 | 2443 |
| | S | 2320 | 0 | 0 | 0 | 0 | 0 | 0 | 0 | 0 | 0 | 0 |
| | T | 2443 | 2443 | 2443 | 2443 | 2443 | 2443 | 2443 | 2443 | 2443 | 2443 | 2443 |
| Decreased clearance | R | 122 | 2443 | 2443 | 2443 | 2443 | 2443 | 2443 | 2443 | 2443 | 2443 | 2443 |
| | S | 2320 | 0 | 0 | 0 | 0 | 0 | 0 | 0 | 0 | 0 | 0 |
| | T | 2443 | 2443 | 2443 | 2443 | 2443 | 2443 | 2443 | 2443 | 2443 | 2443 | 2443 |
| Plasmid acquisition | R | 122 | 2443 | 2443 | 2443 | 2443 | 2443 | 2443 | 2443 | 2443 | 2443 | 2443 |
| | S | 2320 | 0 | 0 | 0 | 0 | 0 | 0 | 0 | 0 | 0 | 0 |
| | T | 2443 | 2443 | 2443 | 2443 | 2443 | 2443 | 2443 | 2443 | 2443 | 2443 | 2443 |
| Antibiotic use, 50% clearance | R | 122 | 2443 | 2443 | 2443 | 2443 | 2443 | 2443 | 2443 | 2443 | 2443 | 2443 |
| | S | 2320 | 0 | 0 | 0 | 0 | 0 | 0 | 0 | 0 | 0 | 0 |
| | T | 2443 | 2443 | 2443 | 2443 | 2443 | 2443 | 2443 | 2443 | 2443 | 2443 | 2443 |
| Antibiotic use in hospital only | R | 122 | 2443 | 2443 | 2443 | 2443 | 2443 | 2443 | 2443 | 2443 | 2443 | 2443 |
| | S | 2320 | 0 | 0 | 0 | 0 | 0 | 0 | 0 | 0 | 0 | 0 |
| | T | 2443 | 2443 | 2443 | 2443 | 2443 | 2443 | 2443 | 2443 | 2443 | 2443 | 2443 |

*R = resistant, S = susceptible, T= total*

**Table S8.** Observed growth reported in Ammerlaan et al. (4) and expected growth

| Observed | 1998 | 2007 |
|---|---|---|
| ARB Enterobacteriaceae | 0·6 | 10·2 |
| ABS Enterobacteriaceae | 22·0 | 35·7 |
| Total | 22·6 | 45·9 |
| Expected | | |
| ARB Enterobacteriaceae | 0·6 | 0·97 |
| ABS Enterobacteriaceae | 22·0 | 35·7 |
| Total | 22·6 | 36·67 |
